# Neurocognitive Benefit of Weight-Loss Interventions in Older Patients with Heart Failure with Preserved Ejection Fraction

**DOI:** 10.1101/2022.11.13.22282230

**Authors:** Christopher L. Schaich, Timothy M. Hughes, Youngkyoo Jung, Dalane W. Kitzman, Haiying Chen, Barbara J. Nicklas, Denise K. Houston, Peter Brubaker, Anthony J.A. Molina, Christina E. Hugenschmidt

**Affiliations:** Department of Surgery, Hypertension and Vascular Research Center, Wake Forest University School of Medicine, Winston-Salem, NC; Department of Internal Medicine, Section on Gerontology and Geriatric Medicine, Wake Forest University School of Medicine; Department of Epidemiology and Prevention, Wake Forest University School of Medicine; Department of Radiology, UC Davis School of Medicine, Sacramento, CA; Department of Internal Medicine, Section on Cardiovascular Medicine, Wake Forest University School of Medicine; Department of Biostatistics and Data Science, Wake Forest University School of Medicine; Department of Health and Exercise Science, Wake Forest University, Winston-Salem, NC; Division of Geriatrics, Gerontology and Palliative Care, UC San Diego School of Medicine, La Jolla, CA

**Keywords:** Heart failure with preserved ejection fraction, cognition, brain, brain imaging, cerebral blood flow, obesity, aging

## Abstract

**Objectives:** Evaluate neurocognitive health and its response to interventions in older, obese patients with heart failure with preserved ejection fraction (HFpEF).

**Background:** Neurocognitive dysfunction may be an underrecognized feature of HFpEF that responds to weight-loss interventions.

**Methods:** We first compared detailed baseline cognitive testing (Uniform Data Set version 3 and Rey Auditory Verbal Learning Test [RAVLT]), and brain volumes and cerebral blood flow (CBF) from 3T magnetic resonance imaging between older patients with HFpEF (n=46) and healthy age-matched controls (HC; n=22). The HFpEF patients were then randomized to a 20-week caloric restriction (CR) intervention with either aerobic-only (CR+AT; n=23) or aerobic+resistance exercise training (CR+AT+RT; n=23), and repeated cognitive testing and neuroimaging post-intervention. Cognitive scores were normalized to national data and transformed to *z*-scores for global, memory, attention, executive function, visuospatial, and language fluency domains.

**Results:** Compared to HC, participants with HFpEF had significantly lower baseline global cognitive performance, and lower global, visuospatial processing and language fluency domain *z*-scores than normative means. Following the diet and exercise intervention, there were significant improvements in global (+0.6 [95% CI: 0.3, 0.8]) and category fluency (+0.2 [95% CI: −0.004, 0.3]) *z*-scores, and in RAVLT immediate (+0.6 [95% CI: 0.1, 1.0] points) and delayed (0.9 [95% CI: 0.2, 1.6] points) recall. Only CR+AT+RT was associated with improved phonemic fluency z-score (+0.4 [95% CI: 0.1, 0.7]). There were no significant intervention effects on brain volumes or CBF.

**Conclusions:** Older, obese patients with chronic HFpEF have significant cognitive deficits that are ameliorated by diet and exercise interventions.

## INTRODUCTION

Heart failure (HF) with preserved ejection fraction (HFpEF) is the most common form of HF in the United States, particularly in older persons and women, and is associated with high morbidity and mortality.^1^ The primary manifestation of HFpEF is severe exercise intolerance, experienced as exertional dyspnea and fatigue, which contributes to patients’ significantly reduced quality of life.^2,3^ In addition to cardiac dysfunction, extra-cardiac factors contributing to exercise intolerance in HFpEF include arterial stiffness, endothelial and mitochondrial dysfunction, capillary rarefaction, and reduced O_2_ extraction in skeletal muscle and myocardium.^3–6^ Exercise training and caloric restriction (CR), first reported by our group, were the first interventions shown to improve exercise capacity and quality of life among HFpEF patients,^7,8^ and appear to exert benefit primarily through extra-cardiac mechanisms.^3,9^

The current paradigm of HFpEF suggests it is a systemic disorder associated with multiple comorbidities, particularly overweight/obesity (present in up to 85% of HFpEF patients), and with increased inflammation that adversely affects most organ systems.^10^ Other common comorbidities of HFpEF include hypertension, dysglycemia, and dyslipidemia, which impair vascular function and contribute to its pathophysiology.^11^ These comorbidities are each consistently and synergistically associated with cognitive decline, brain structural abnormalities, and greater risk for all-cause dementia.^12,13^ Given its systemic vascular pathology and the highly vascularized nature of the brain, cognitive dysfunction may be a common but underrecognized feature of HFpEF. Indeed, the more widely-studied HF with reduced ejection fraction (HFrEF) is associated with progressive cognitive decline and increased risk for dementia, potentially via alterations to cerebral blood flow (CBF).^14,15^

However, comparatively little is known regarding neurocognitive effects of HFpEF. Additionally, CR and exercise interventions have been shown to have a beneficial role in preserving brain health and cognition during aging and in persons with obesity.^16–18^ However, no studies have systematically examined detailed cognitive function, brain structure, and CBF in HFpEF patients compared to healthy age-matched controls, nor assessed the impact of CR and exercise on cognitive function in HFpEF. Therefore, our objective was to fill these important knowledge gaps.

## METHODS

### Parent trial design

This analysis was ancillary to the Study of the Effect of Caloric Restriction and Exercise Training in Patients with Heart Failure and a Normal Ejection Fraction (SECRET-II) (ClinicalTrials.gov identifier NCT02636439), a randomized, single-blind, parallel assignment clinical trial in 88 men and women with HFpEF conducted at Wake Forest University School of Medicine (Winston-Salem, NC) from August 2015 to July 2021. Its objective was to determine if the addition of resistance training (RT) to a 20-week weight loss intervention of CR and aerobic exercise training (AT) improved peak exercise oxygen consumption (VO_2_) in older adults with stable HFpEF. Participants were randomized to CR+AT or CR+AT+RT interventions. In accord with prior published trials of older obese HFpEF,^8^ eligibility criteria were: age ≥60 years; body mass index (BMI) ≥28 kg/m^2^; National Health and Nutrition Examination Survey Congestive HF Clinical Score ≥3 or the Rich criteria;^19^ left ventricular ejection fraction ≥50%; and diastolic dysfunction ≥grade 1. Exclusion criteria are listed in Supplemental Table 1. The trial was approved by the Wake Forest University School of Medicine Institutional Review Board and completed in accordance with the Declaration of Helsinki. All participants provided written informed consent prior to enrollment.

### Participants

All SECRET-II trial participants enrolled after July 13, 2017 (N=46; 23 in each intervention arm) completed, in addition to other trial assessments, a baseline and post-intervention cognitive assessment. Of these, 43 completed baseline brain magnetic resonance imaging (MRI) and 40 completed post-intervention brain MRI. A group of age- and sex-matched participants (n=22) with no significant chronic medical conditions were recruited to serve as controls for comparison and underwent identical baseline assessments. Healthy controls were excluded if they were on chronic medications other than preventive low-dose aspirin, had an abnormal physical examination (including hypertension) or abnormal screening test results (including electrocardiogram, exercise echocardiogram, and spirometry), or regularly undertook vigorous exercise.

### Clinical and physical measures

Participants self-reported their age, sex, race/ethnicity, years of education, and history of diabetes diagnosis or medication use. Hypertension was defined as systolic blood pressure ≥130 mmHg or taking antihypertensive medication. BMI (kg/m^2^) was calculated from measured height and weight. VO_2_ (ml/kg/min) was measured during a progressive exercise test on a motorized treadmill using the modified Naughton protocol to the endpoint of exhaustion (respiratory exchange ratio >1.05 or >90% age-predicted maximal heart rate).^20^ Expired gases were measured with the Ultima system (MedGraphics, Minneapolis, MN) following calibration. Peak VO_2_ was defined as the average of the final 45 seconds of peak exercise. Participants also completed the Short Physical Performance Battery^21^ (SPPB) to assess gait speed over 4 meters, timed repeated chair stand, and standing balance. Scores on each component were summed to provide a score of range 0-12, with higher scores indicating better physical performance.

### Evaluation of cognitive function

Cognitive testing was conducted in a quiet room by a trained administrator at the baseline and post-intervention visits. Participants completed the Rey Auditory Verbal Learning Test (RAVLT) and the Uniform Data Set version 3 (UDSv3) cognitive battery. The RAVLT is a word list memory task,^22^ scored as the sum of correctly recalled words immediately following the task and after a 25-min delay. The UDSv3 is a standardized cognitive battery used by all NIH-funded Alzheimer’s Disease Centers^23^ and includes the Montreal Cognitive Assessment (MoCA), a test of global cognitive function. Other UDSv3 tests included Craft Story (immediate and delayed recall verbatim and paraphrased); Benson Complex Figure (copy and recall); phonemic fluency test (letters F and L); category fluency (Animals and Vegetables); Number Span forward and backward (total correct and longest span); and Trail Making Test A and B. Raw scores on the MoCA and other UDSv3 tests were normalized against national population data to create *z*-scores based on age, sex, race, and education,^24^ and combined into domain-specific *z*-scores for memory (Craft Story immediate and delayed recall, Benson Complex Figure recall), attention (Number Span forward and backward), executive function (Trail Making Test), visuospatial processing (Benson Complex Figure copy), and language (category fluency and phonemic fluency).^23^

### Neuroimaging

Brain morphology and CBF were computed from MRI acquired on a 3T Skyra scanner (Siemens, Erlangen, Germany) with a 32-channel head coil. Sequences included T1-weighted imaging, T2-weighted fluid-attenuated inversion recovery (T2-FLAIR), and pseudocontinuous arterial spin labeling (pcASL). Total gray and white matter volume, including frontal and temporal lobe gray matter and hippocampus volumes, were calculated from T1-weighted images using the Computational Anatomy Toolbox (http://www.neuro.uni-jena.de/cat/) in SPM12 (www.fil.ion.ucl.ac.uk/spm) and expressed relative to total intracranial volume (ICV). White matter hyperintensity (WMH) lesion volume relative to ICV was computed by a growth algorithm^25^ using T2-FLAIR images, and its distribution normalized by log transformation. Gray matter CBF was quantified from pcASL images with a kinetic model.^26^ Perfusion-weighted images were converted to physiological units (ml/100 g tissue/min; fully-relaxed cerebrospinal fluid magnetization reference). Gray matter regions of interest were overlaid on CBF maps to calculate mean global, frontal lobe, temporal lobe, and hippocampal CBF. Detailed MRI processing information is provided in the Supplemental Methods.

### Interventions

SECRET-II participants completed a 20-week intervention of CR with exercise training (CR+AT or CR+AT+RT) in accordance with guidelines for adults with overweight/obesity.

#### Caloric Restriction

All participants were prescribed a hypocaloric diet with lunch and dinner plus snacks provided by the Wake Forest Clinical Research Unit Metabolic Kitchen; participants prepared their own breakfast from a menu of recommendations. Participants kept a food log and attended weekly weigh-in sessions that included individual counseling with a registered dietitian. The prescribed calorie level was targeted to achieve a 300 kcal/day deficit, yielding an average daily intake of 1650 kcal/day. Meals were tailored to provide a balanced diet with <30% of calories from fat and 1.3-1.5 g protein/kg/day. No participant received less than 1100 kcal/day for women or 1300 kcal/day for men, and total weight loss was capped at 15% body weight or a BMI of 25 kg/m^2^.

#### Exercise training

Participants exercised 3 times/week in 50-minute supervised sessions. The exercise interventions were individualized and increased in intensity based on specific milestones. Following a 30-minute AT session that included a walking circuit and/or stationary cycling, CR+AT participants completed 20 minutes of light chair-based stretching and flexibility maneuvers, while the CR+AT+RT group completed 20 minutes of RT on Nautilus weight machines. A detailed description of exercise interventions is provided in the Supplemental Methods.

### Statistical analysis

Analyses were completed in R 4.1.2 using a two-tailed significance threshold of *p*<0.05. First, we compared baseline characteristics of participants with HFpEF and healthy controls using two-sample t-test for normally distributed continuous data, Wilcoxon rank-sum test for ordinal data, and Fisher’s exact test for proportions. We then compared baseline MoCA and RAVLT scores and neuroimaging outcomes of participants with HFpEF to controls using analysis of covariance adjusted for age, sex, and race. MoCA and RAVLT scores were additionally adjusted for years of education. Next, we compared the baseline MoCA and cognitive domain *z*-scores of the combined SECRET-II intervention groups against national normative population scores (μ=0, σ=1 by definition) using one-sample t-test to determine if HFpEF patients performed lower than age-, sex-, race-, and education-specific population means in any domain. To assess intervention effects on cognitive performance, brain volumetrics, and CBF we used linear mixed effects models with random intercepts. Models included fixed visit (baseline and post-intervention), intervention (CR+AT and CR+AT+RT), and group-by-visit interaction effects. Analyses of raw MoCA and RAVLT scores included further adjustments for age, sex, race, and years of education; cognitive domain and MoCA *z*-scores were normed to these variables and not adjusted further. Brain volumetrics and CBF were adjusted for age, sex, and race. We report combined and group-specific changes in outcomes from baseline with 95% confidence intervals (CI), and the group-by-visit interaction *p*-value to illustrate randomization effects where appropriate.

## RESULTS

Baseline characteristics, raw MoCA and RAVLT scores, and brain volumetrics and CBF in participants with HFpEF and healthy controls are provided in Table 1. Participants with HFpEF had a median [25%, 75%] baseline MoCA score of 24 [23, 26] out of 30, and performed significantly worse than healthy controls after adjustment for age, sex, race, and years of education (*p*=0.017); MoCA scores under 26 and above 17 indicate mild cognitive impairment (mocatest.org/faq/). Baseline characteristics of participants with HFpEF stratified by intervention group are shown in Supplemental Table 2. Participants randomized to CR+AT+RT tended to be older (70.2 [5.7] vs. 67.2 [4.8] years; *p*=0.06), but there were no other group differences in baseline characteristics. Unadjusted MoCA and RAVLT scores, and normed MoCA and cognitive domain *z*-scores among participants with HFpEF at baseline and post-intervention are shown in Table 2. There were no between-group differences in baseline cognitive scores (all *p* >0.10). Participants with HFpEF scored significantly lower than normative population means on the MoCA and in visuospatial processing, category fluency, and phonemic fluency domains (Table 2).

**Table 1.**
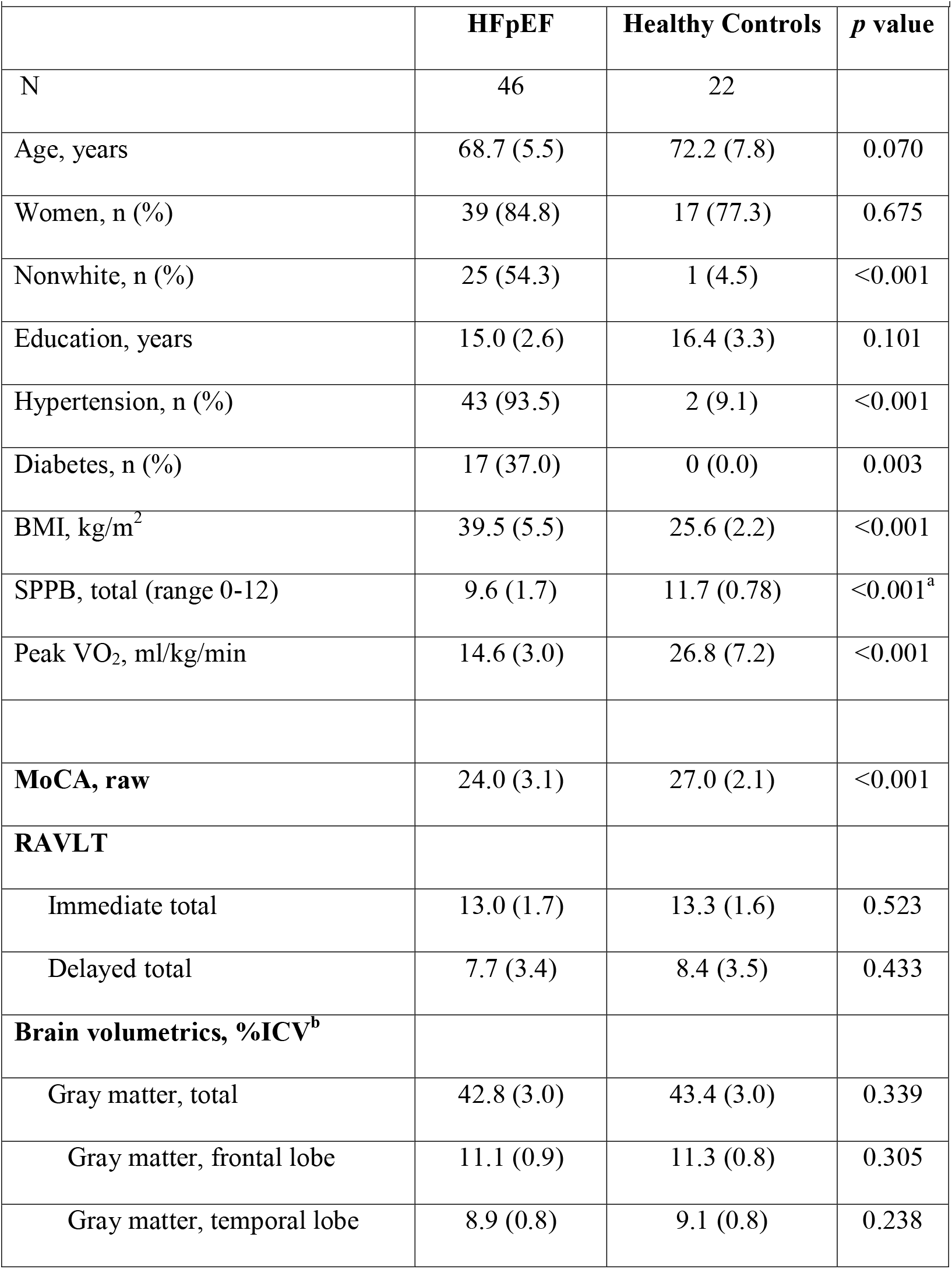
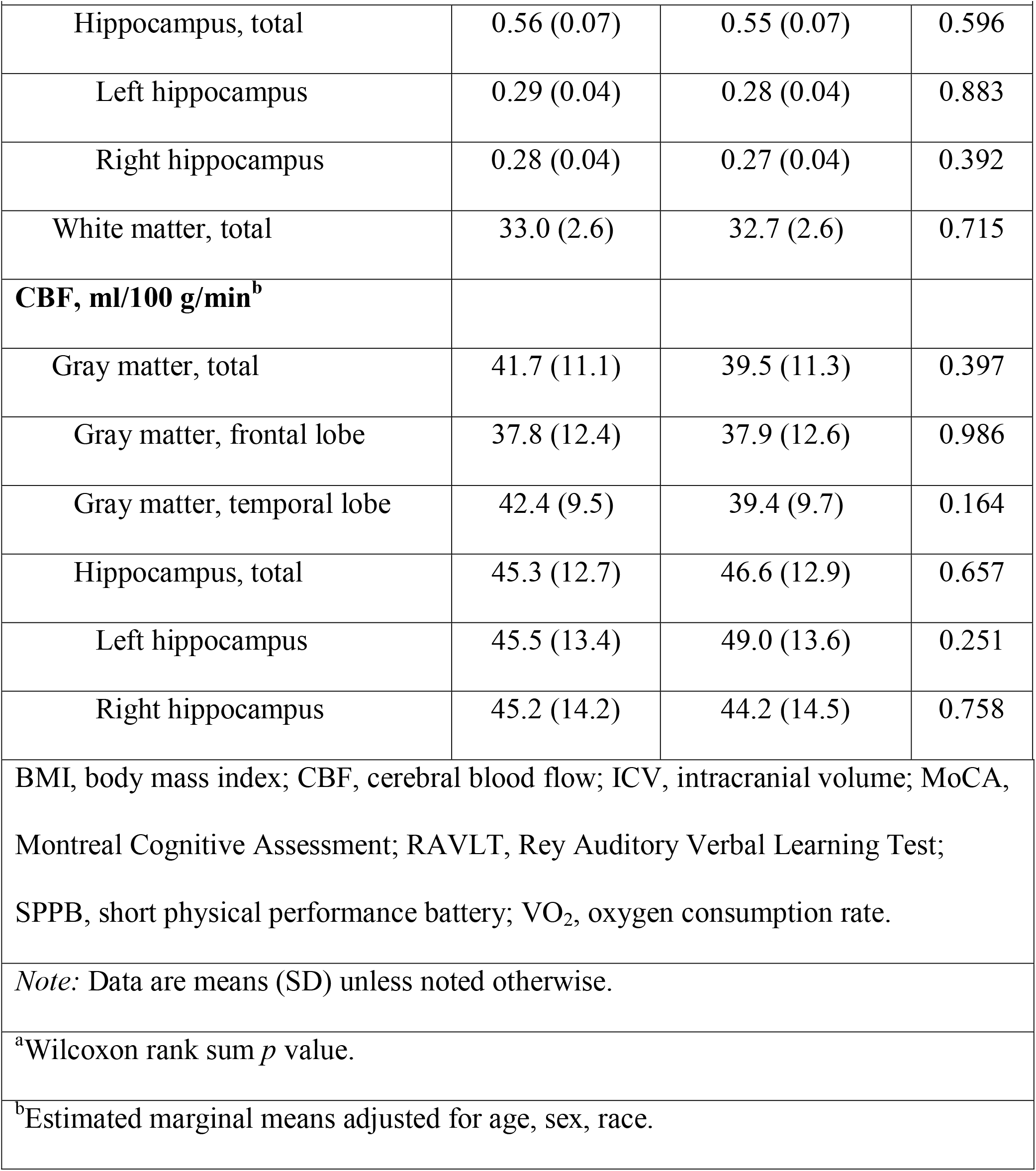
Comparison of baseline characteristics, cognitive function, brain volumetrics, and cerebral blood flow in participants with HFpEF and age- and sex-matched controls.

**Table 2.**
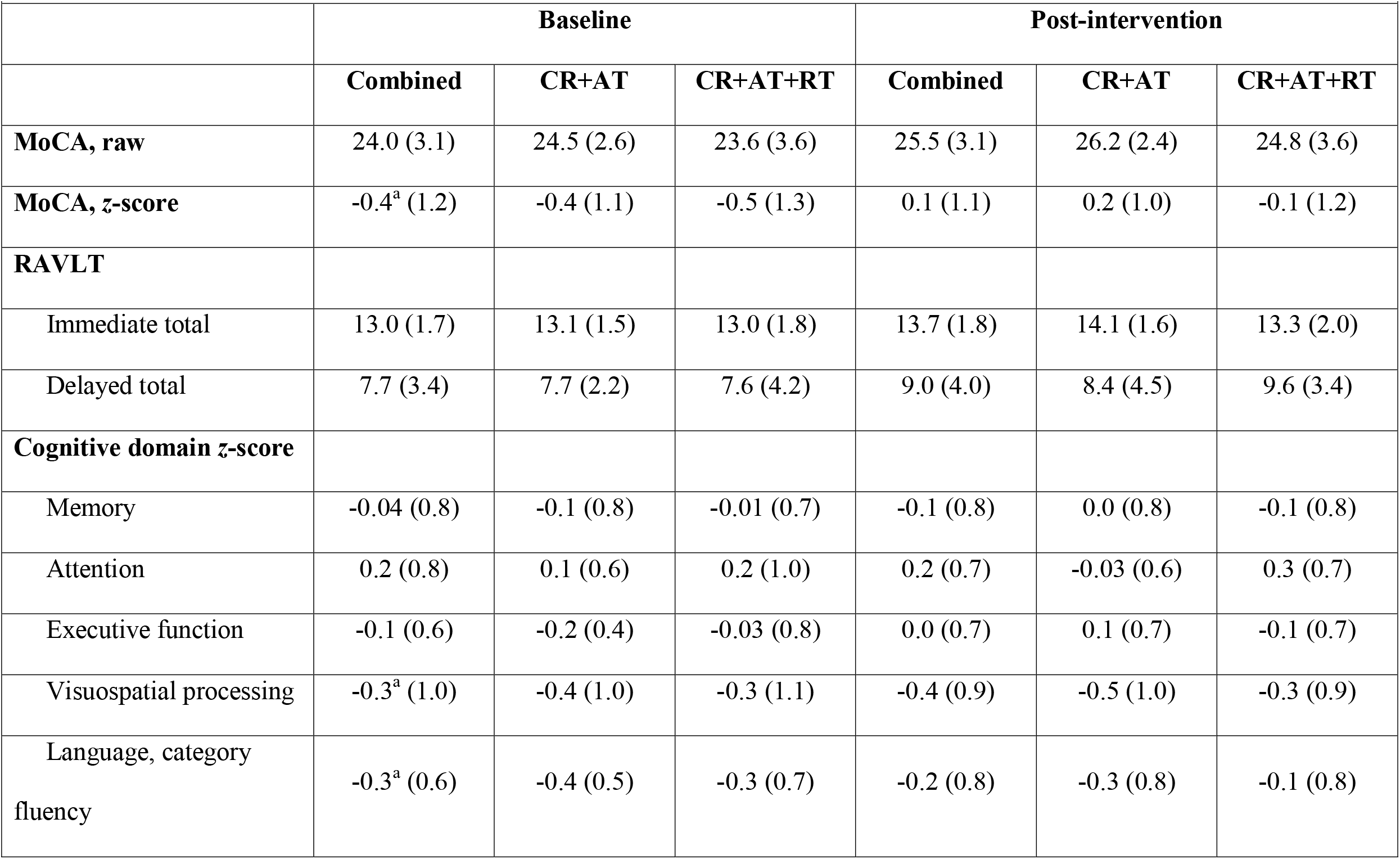
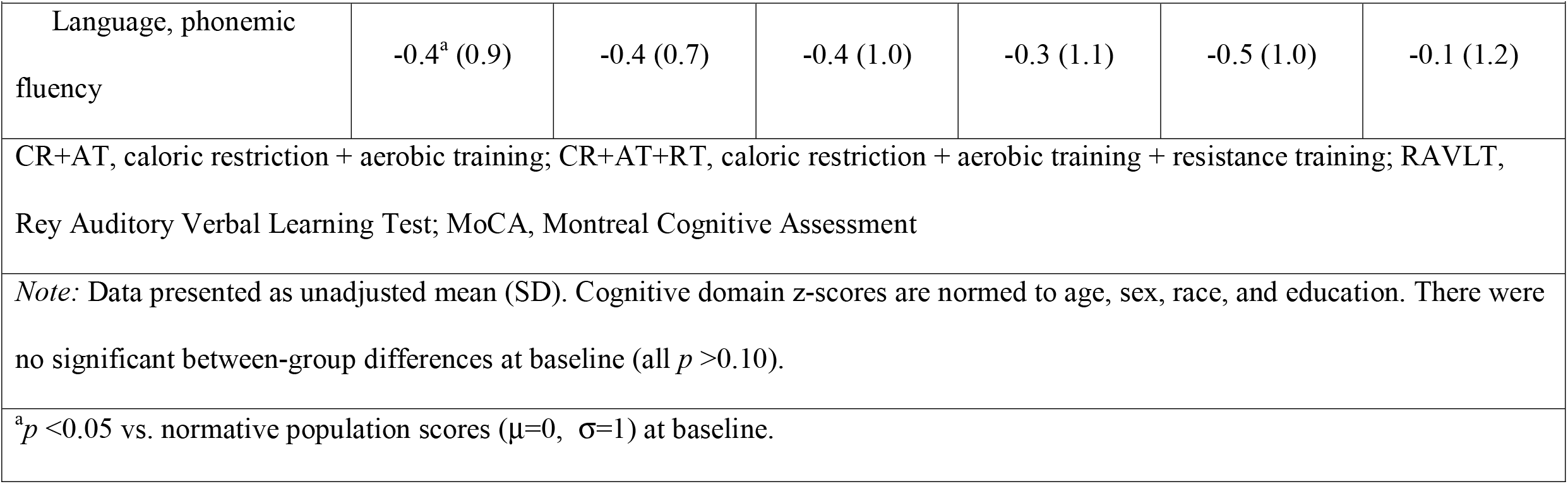
Unadjusted mean cognitive outcome scores before and after caloric restriction and exercise interventions.

|  | **Baseline** |  |  | **Post-intervention** |  |  |
| --- | --- | --- | --- | --- | --- | --- |
|  | **Combined** | **CR+AT** | **CR+AT+RT** | **Combined** | **CR+AT** | **CR+AT+RT** |
| **MoCA, raw** | 24.0 (3.1) | 24.5 (2.6) | 23.6 (3.6) | 25.5 (3.1) | 26.2 (2.4) | 24.8 (3.6) |
| **MoCA, z-score** | -0.4<sup>a</sup> (1.2) | -0.4 (1.1) | -0.5 (1.3) | 0.1 (1.1) | 0.2 (1.0) | -0.1 (1.2) |
| **RAVLT** |  |  |  |  |  |  |
| Immediate total | 13.0 (1.7) | 13.1 (1.5) | 13.0 (1.8) | 13.7 (1.8) | 14.1 (1.6) | 13.3 (2.0) |
| Delayed total | 7.7 (3.4) | 7.7 (2.2) | 7.6 (4.2) | 9.0 (4.0) | 8.4 (4.5) | 9.6 (3.4) |
| **Cognitive domain z-score** |  |  |  |  |  |  |
| Memory | -0.04 (0.8) | -0.1 (0.8) | -0.01 (0.7) | -0.1 (0.8) | 0.0 (0.8) | -0.1 (0.8) |
| Attention | 0.2 (0.8) | 0.1 (0.6) | 0.2 (1.0) | 0.2 (0.7) | -0.03 (0.6) | 0.3 (0.7) |
| Executive function | -0.1 (0.6) | -0.2 (0.4) | -0.03 (0.8) | 0.0 (0.7) | 0.1 (0.7) | -0.1 (0.7) |
| Visuospatial processing | -0.3<sup>a</sup> (1.0) | -0.4 (1.0) | -0.3 (1.1) | -0.4 (0.9) | -0.5 (1.0) | -0.3 (0.9) |
| Language, category<br>fluency | -0.3<sup>a</sup> (0.6) | -0.4 (0.5) | -0.3 (0.7) | -0.2 (0.8) | -0.3 (0.8) | -0.1 (0.8) |
| Language, phonemic fluency | -0.4<sup>a</sup> (0.9) | -0.4 (0.7) | -0.4 (1.0) | -0.3 (1.1) | -0.5 (1.0) | -0.1 (1.2) |
| CR+AT, caloric restriction + aerobic training; CR+AT+RT, caloric restriction + aerobic training + resistance training; RAVLT, Rey Auditory Verbal Learning Test; MoCA, Montreal Cognitive Assessment<br>*Note:* Data presented as unadjusted mean (SD). Cognitive domain z-scores are normed to age, sex, race, and education. There were no significant between-group differences at baseline (all $p > 0.10$).<br><sup>a</sup>$p < 0.05$ vs. normative population scores ($\mu=0$, $\sigma=1$) at baseline. |  |  |  |  |  |  |

### Overall intervention effects

Table 3 displays intervention effects on cognitive function in participants with HFpEF. Performance in the overall sample improved on the MoCA (+1.46 [95% CI: 0.83, 2.08] points vs. baseline; +0.56 [95% CI: 0.31, 0.81] *z*-score vs. baseline), and on the immediate (+0.55 [95% CI: 0.05, 1.04] words vs. baseline) and delayed recall (+0.87 [95% CI: 0.16, 1.59] words vs. baseline) portions of the RAVLT (Table 3). There was limited improvement in the language domains (category fluency, +0.15 [95% CI: −0.004, 0.31] *z*-score; phonemic fluency, +0.18 [95% CI: −0.03, 0.39] *z*-score), but not in other domains. These effects were similar after additional adjustments for changes in BMI, VO_2_, total gray matter volume, or gray matter CBF (Supplemental Table 3).

**Table 3.** Effect of caloric restriction and exercise interventions on cognitive outcomes from adjusted linear mixed effects models.

|  |  |  |  |
| --- | --- | --- | --- |
|  | **Change from baseline (95% CI)** |  |  |
|  | **Combined** | **CR+AT** | **CR+AT+RT** |
| **MoCA, raw<sup>a</sup>** | 1.46 (0.83, 2.08) | 1.68 (0.77, 2.59) | 1.24 (0.39, 2.09) |
| **MoCA, z-score** | 0.56 (0.31, 0.81) | 0.65 (0.28, 1.01) | 0.48 (0.14, 0.82) |
| **RAVLT<sup>a</sup>** |  |  |  |
| Immediate total | 0.55 (0.05, 1.04) | 0.90 (0.18, 1.63) | 0.19 (-0.48, 0.86) |
| Delayed total | 0.87 (0.16, 1.59) | 1.33 (0.28, 2.38) | 0.42 (-0.56, 1.39) |
| **Cognitive domain z-score** |  |  |  |
| Memory | -0.005 (-0.20, 0.19) | 0.10 (-0.18, 0.39) | -0.11 (-0.38, 0.16) |
| Attention | 0.03 (-0.15, 0.20) | -0.06 (-0.31, 0.20) | 0.11 (-0.14, 0.35) |
| Executive function | 0.11 (-0.11, 0.33) | 0.25 (-0.07, 0.58) | -0.04 (-0.34, 0.27) |
| Visuospatial | -0.04 (-0.43, 0.35) | -0.11 (-0.68, 0.45) | 0.03 (-0.50, 0.57) |
| Language, category<br>fluency | 0.15 (-0.004, 0.31) | 0.12 (-0.11, 0.35) | 0.18 (-0.03, 0.40) |

| Table 3, continued. |  |  |  |
| --- | --- | --- | --- |
| Language, phonemic fluency | 0.18 (-0.03, 0.39) | -0.03 (-0.34, 0.28) | 0.38 (0.09, 0.68) |
| CR+AT, caloric restriction+aerobic training; CR+AT+RT, caloric restriction+aerobic training+resistance training; RAVLT, Rey Auditory Verbal Learning Test; MoCA, Montreal Cognitive Assessment |  |  |  |
| <sup>a</sup>Adjusted for age, sex, race, and years of education. *Note:* MoCA and cognitive domain z-scores are normed to these values. |  |  |  |

### Effect of randomization

Randomization to CR+AT or CR+AT+RT did not produce significant between-group differences in changes in MoCA or RAVLT scores (all interaction *p* >0.10). However, only the CR+AT intervention was associated with significant improvement on the immediate and delayed recall portions of the RAVLT after adjustment for age, sex, race, and years of education (Table 3). In contrast, only randomization to CR+AT+RT was associated with improvement in the phonemic fluency domain (CR+AT: −0.03 [95% CI: −0.38, 0.32] vs. CR+AT+RT: +0.38 [95% CI: 0.09, 0.68] SD units; interaction *p*=0.06). There were no combined or group-specific changes in total or regional brain volumes or CBF after the 20-week intervention (Table 4).

**Table 4.** Caloric restriction and exercise intervention effects on brain volumetrics and CBF.

|  |  |  |  |
| --- | --- | --- | --- |
|  | **Change from baseline (95% CI)<sup>a</sup>** |  |  |
|  | **Combined** | **CR+AT** | **CR+AT+RT** |
| **Brain volumetrics, %ICV** |  |  |  |
| Gray matter, total | 0.19 (-0.25, 0.62) | 0.31 (-0.31, 0.93) | 0.06 (-0.56, 0.68) |
| Gray matter, frontal lobe | 0.02 (-0.10, 0.13) | 0.01 (-0.16, 0.18) | 0.03 (-0.14, 0.19) |
| Gray matter, temporal lobe | 0.08 (-0.07, 0.22) | 0.13 (-0.07, 0.33) | 0.02 (-0.18, 0.22) |
| Hippocampus, total | 0.003 (-0.004, 0.01) | 0.01 (-0.004, 0.02) | 0.001 (-0.01, 0.01) |
| Left hippocampus | 0.003 (-0.002, 0.007) | 0.004 (-0.003, 0.01) | 0.002 (-0.005, 0.008) |
| Right hippocampus | 0.001 (-0.002, 0.004) | 0.003 (-0.002, 0.007) | -0.001 (-0.01, 0.004) |
| White matter, total | 0.04 (-0.16, 0.24) | -0.03 (-0.31, 0.26) | 0.11 (-0.18, 0.40) |
| **White Matter Hyperintensity,**<br>**log(%ICV)** | -0.002 (-0.02, 0.01) | -0.004 (-0.02, 0.02) | -0.001 (-0.02, 0.02) |
| **CBF, ml/100 g tissue/min** |  |  |  |
| Gray matter, total | -0.78 (-3.73, 2.17) | -2.24 (-6.40, 1.91) | 0.69 (-3.50, 4.88) |
| Gray matter, frontal lobe | -0.40 (-3.74, 2.95) | -2.15 (-6.85, 2.55) | 1.36 (-3.39, 6.11) |

| **Table 4, continued.** |  |  |  |
| --- | --- | --- | --- |
| Gray matter, temporal lobe | -0.11 (-2.73, 2.51) | -0.67 (-4.36, 3.01) | 0.46 (-3.26, 4.17) |
| Hippocampus, total | 1.22 (-1.83, 4.27) | -0.04 (-4.34, 4.27) | 2.47 (-1.86, 6.80) |
| Left hippocampus | 0.30 (-3.12, 3.71) | -1.47 (-6.27, 3.34) | 2.06 (-2.79, 6.91) |
| Right hippocampus | 2.13 (-1.53, 5.78) | 1.30 (-3.85, 6.45) | 2.95 (-2.23, 8.14) |
| CR+AT, caloric restriction + aerobic training intervention; CR+AT+RT, caloric restriction + aerobic training + resistance training intervention; CBF, cerebral blood flow; ICV, total intracranial volume. |  |  |  |
| <sup>a</sup>Adjusted for age, sex, and race. |  |  |  |

## DISCUSSION

In this cross-sectional comparison followed by a randomized clinical trial ancillary study, we found that older adults with chronic, stable HFpEF had worse global cognition relative to nationally representative population data and a healthy adult control sample, with domain-specific deficits in language (category fluency and phonemic fluency) and visuospatial processing. Global cognitive performance and word list learning and memory recall improved significantly in the HFpEF patients after completing a 20-week CR+exercise intervention. There was some evidence of improvement in language domains, though mostly in the CR+AT+RT group. These improvements were independent of cerebral gray matter volume and CBF, which were unchanged following the interventions. We believe these results provide the first evidence that global and domain-specific cognitive deficits in chronic, stable HFpEF may be ameliorated by CR+exercise training.

Cognitive dysfunction could be a major contributor to worse outcomes in HF by interfering with disease management or patient safety (e.g. poor medication compliance, missed appointments, or worse self-care).^27^ However, to date, most literature on HF-related neurocognitive outcomes is in patients with HFrEF or undifferentiated HF (see Vishwanath et al. [2021]^28^ for recent systematic review and meta-analysis). In particular, HFrEF is associated with increased risk for vascular dementia^29^ and Alzheimer’s disease.^30^ Moreover, low ejection fraction and subclinical changes in cardiac output and stroke volume were found to be associated with slower processing speed, worse executive function, lower brain volumes, and higher risk for dementia.^31,32^ In contrast, there have been few previous neuroimaging studies in HFpEF, and we are unaware of any that compared brain volumes or CBF. We believe our study is the first to investigate detailed cognitive function, brain volumes, and CBF in chronic, stable HFpEF, and suggests that significant global impairment is present in these patients as well.

Cerebral hypoperfusion precipitates cognitive decline,^33^ and there is evidence that HF may result in chronic cerebral hypoperfusion globally and in key regions for cognition.^14,34^ Moreover, changes in CBF in HFrEF patients are associated with structural brain abnormalities including reduced gray matter and increased WMH volume.^35,36^ In HFpEF, where systemic capillary rarefaction is a key pathologic finding,^5,6^ similar capillary pruning in the brain could contribute to hypoperfusion with consequent cognitive impairment. However, we found no differences in age-, sex-, and race-adjusted CBF or brain volumetrics between our sample of HFpEF patients and healthy older adult controls. Furthermore, the observed improvement in global cognitive performance following the intervention was independent of CBF, which did not change significantly. Thus, our results suggest that mechanisms other than chronic cerebral hypoperfusion account for the significant cognitive impairment we observed in participants with HFpEF.

Both CR and AT have previously been shown to significantly improve exercise intolerance in HFpEF.^7,8^ Of note, these interventions have also shown benefit to the brain and cognition in sedentary older adults.^16–18^ A growing body of evidence also suggests RT benefits cognitive function in healthy older adults and in adults with mild cognitive impairment.^37–39^ The effects of combined AT+RT on cognition are not well studied, though small randomized trials have found benefit after stroke^40^ and in men with COPD^41^ compared to either training alone. In our study, the CR+AT+RT group showed evidence of greater improvement in language domains compared to the CR+AT group, despite similar BMI and exercise capacity at baseline. Further study is warranted to clarify the effect of combined exercise on neurocognitive function in HFpEF.

The mechanisms of exercise-related benefits for brain health are unclear, but could involve integrative effects on multiple components including improved cardiorespiratory fitness, reduced systemic inflammation, or improved endothelial or mitochondrial function.^8,42^ Enhanced cerebrovascular function is another plausible mechanism, as aerobic exercise in mice was associated with increased angiogenesis in the brain.^43^ Improved O_2_ extraction, observed in thigh muscle and the frontal lobe of HFpEF but not HFrEF patients following AT,^44^ is another potential mechanism. These findings are consistent with evidence that CR and exercise appear to exert benefit in HFpEF through improved systemic microvascular function.^9^ However, further study is needed to determine the mechanisms of exercise training benefits to cognition in HFpEF.

### Strengths and limitations

This study has several strengths, including prospective design, a detailed cognitive assessment battery, concomitant brain imaging, inclusion of well-defined HFpEF and healthy control cohorts, moderate sample size, and application of two non-pharmacologic interventions previously shown to benefit patients with HFpEF. This study also has potential limitations. It was designed to collect longitudinal pilot data on cognition and the brain in HFpEF by adding measures to an ongoing clinical trial. Thus, it was not powered to detect all meaningful effects in cognitive and brain imaging outcomes. Further, the lack of a usual care or CR-only arm complicates the interpretation of effects on cognition and brain imaging outcomes because these have not been characterized previously. We assessed effects immediately following the intervention, while it is possible that exercise benefits to brain health evolve over different intervention and post-intervention durations. Finally, the lack of observed change in CBF could be due to a need for methodological optimization in HFpEF. For example, protocol specifications such as transit time delay (time for tagged blood to perfuse) can vary with age and vascular disorders. More studies are needed to optimize methods for testing cerebrovascular health in people with HFpEF.

## Conclusions

HFpEF is a common geriatric syndrome with systemic, multi-organ involvement and significant vascular and other extra-cardiac contributors that may predispose patients to neurocognitive dysfunction **(Central Illustration)**. Our study supports this hypothesis and, to our knowledge, is the first to assess detailed cognition in chronic, stable HFpEF patients at baseline compared to healthy age-matched controls, and after an intervention. These results provide the first evidence that global and domain-specific cognitive deficits are present in chronic, stable HFpEF, and that they may be ameliorated by CR+exercise interventions. Deeper neurocognitive phenotyping may help elucidate the mechanisms of these impairments at baseline and their improvement, as well as the potential for improvement with other interventions and in other HFpEF phenotypes.

## PERSPECTIVES

### Competency in medical knowledge

This ancillary study found that patients with chronic, stable HFpEF exhibited significant deficits in global cognition and language fluency domains that were alleviated by CR and exercise interventions.

### Translational outlook

Cognitive impairment may be an underrecognized but important feature of HFpEF. More detailed neurocognitive phenotyping is needed to determine the pathologic mechanisms of these impairments, as well as to discover which interventions will provide the most benefit to brain health in patients with HFpEF.

## Data Availability

All data produced in the present study are available upon reasonable request from researchers trained in the responsible conduct of research in human subjects.

## NON-STANDARD ABBREVIATIONS

AT: aerobic exercise training
BMI: body mass index
CBF: cerebral blood flow
CI: confidence interval
CR: caloric restriction
FLAIR: fluid-attenuated inversion recovery
HF: heart failure
HFpEF: heart failure with preserved ejection fraction
HFrEF: heart failure with reduced ejection fraction
ICV: intracranial volume
MRI: magnetic resonance imaging
MoCA: Montreal Cognitive Assessment
pcASL: pseudocontinuous arterial spin labeling
RAVLT: Rey Auditory Verbal Learning Test
RT: resistance exercise training
SECRET-II: Study of the Effect of Caloric Restriction and Exercise Training in Patients with Heart Failure and a Normal Ejection Fraction
UDSv3: Uniform Data Set version 3
WMH: white matter hyperintensity

## ACKNOWLEDGEMENTS

We thank all SECRET-II participants and staff, including Ben Nelson and Russel Newland, for their valuable contributions. We thank the MRI technicians involved with the study, especially Sandy Kaminsky.

## FIGURE LEGENDS

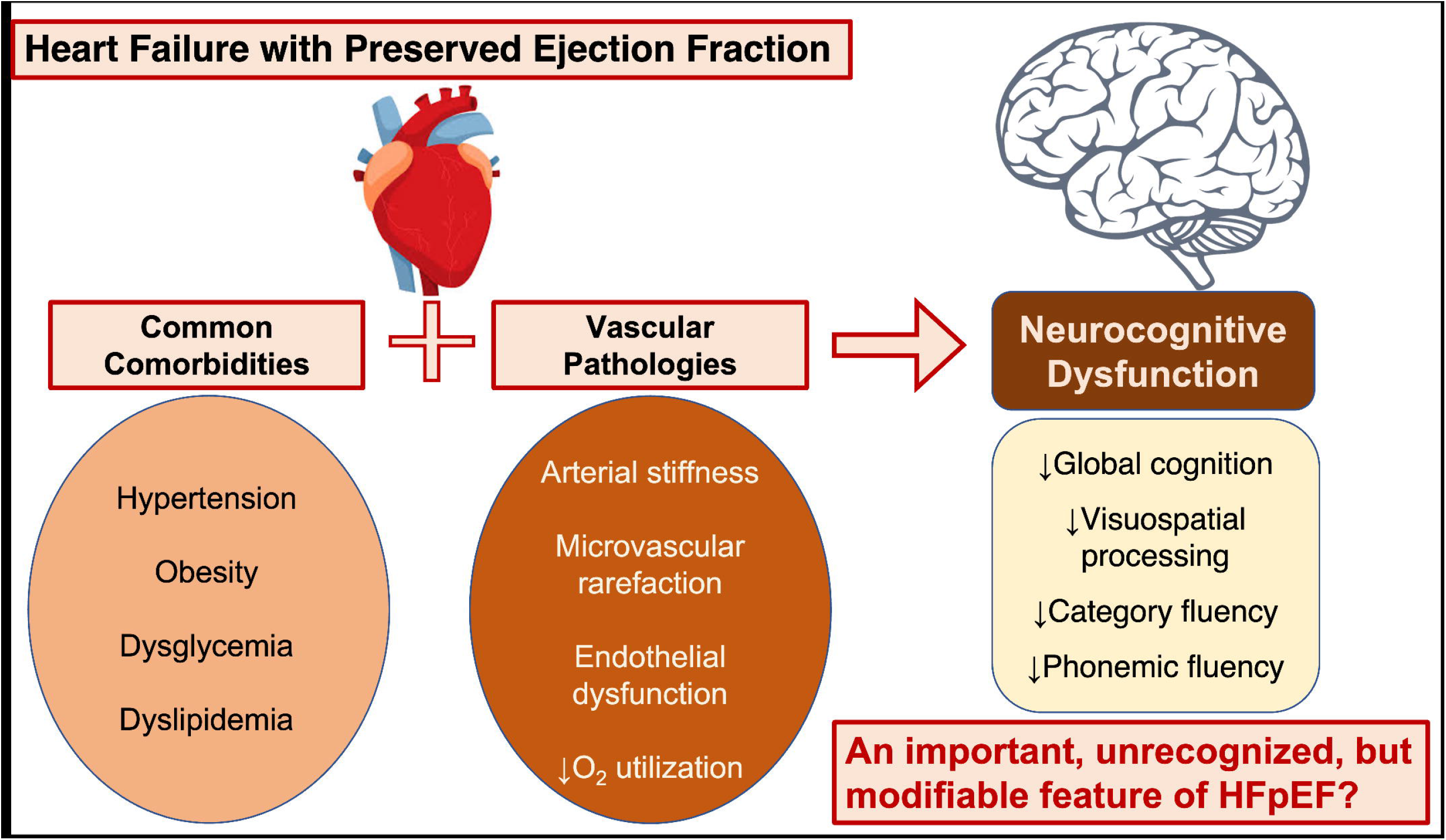
Central Illustration: Common comorbidities and vascular pathologies associated with HFpEF may predispose older patients to neurocognitive dysfunction, which may be alleviated by weight-loss interventions.

