## Supplemental Appendix for "Neurocognitive Benefit of Weight-Loss Interventions in Older Patients with Heart Failure with Preserved Ejection Fraction"

This document contains the following:

Page

**Supplemental Table 1:** Inclusion and exclusion criteria of the SECRET-II trial.	2

**Supplemental Table 2:** Baseline demographic and clinical characteristics of participants with

HFpEF by intervention group.	3

**Supplemental Table 3:** Effect of CR and exercise interventions on cognitive outcomes from

linear mixed effects models additionally adjusted for BMI, peak VO_2_, gray matter volume, or cerebral blood flow at baseline and follow-up.	4

**Supplemental Methods**

Exercise training	9

MRI data processing	11

**Supplemental References**	12

| **Supplemental Table 1.** Inclusion and exclusion criteria of the SECRET-II trial. | |
| --- | --- |
| **Inclusion Criteria** | **Exclusion Criteria** |
| Age ≥60 years | Valvular heart disease as primary etiology of HF |
| BMI ≥28 kg/m^2^ | Change in cardiac medication or HF symptoms, hospitalization or urgent care visit in <6 weeks |
| NHANES HF Clinical Score ≥3 | Uncontrolled hypertension, diabetes, or psychiatric disease (major psychoses, depression, dementia, or personality disorder) |
| LV ejection fraction ≥50% | Evidence of significant COPD |
| LV Diastolic Dysfunction ≥ grade 1 | Recent or debilitating stroke |
|  | Cancer or other non-cardiovascular conditions with life expectancy <2 years |
|  | Significant anemia or renal insufficiency (<10 g/dL Hgb) |
|  | Significant renal insufficiency (eGFR <30 ml/min/1.73 m^2^) |
|  | Contraindication for exercise intervention, thigh muscle biopsy, or MRI |
|  | Plans to leave area within the study period |
|  | Refuses informed consent |

| **Supplemental Table 2.** Baseline demographic and clinical characteristics of participants with HFpEF by intervention group. | | | |
| --- | --- | --- | --- |
|  | **CR+AT** | **CR+AT+RT** | ***p* value** |
|  | n = 23 | n = 23 |  |
| Age, years | 67.2 (4.8) | 70.2 (5.7) | 0.057 |
| Women, n (%) | 19 (82.6) | 20 (87.0) | 0.681 |
| Nonwhite, n (%) | 13 (56.5) | 12 (52.2) | 0.767 |
| Education, years | 15.3 (2.8) | 14.6 (2.3) | 0.342 |
| Hypertension, n (%) | 22 (95.7) | 21 (91.3) | 0.550 |
| Systolic blood pressure, mmHg | 139.9 (13.3) | 136.2 (15.4) | 0.383 |
| Diabetes, n (%) | 7 (30.4) | 10 (43.5) | 0.360 |
| BMI, kg/m^2^ | 39.9 (5.5) | 39.1 (5.7) | 0.615 |
| SPPB, total (range 0-12) | 10.0 (1.2) | 9.2 (2.1) | 0.165^a^ |
| Peak VO_2_, ml/kg/min | 14.0 (2.9) | 15.2 (3.1) | 0.195 |
| CR+AT, caloric restriction + aerobic training; CR+AT+RT, caloric restriction + aerobic training + resistance training; BMI, body mass index; SPPB, short physical performance battery. | | | |
| *Note:* Data are means (SD) unless noted otherwise. | | | |
| ^a^Wilcoxon rank sum test *p* value. | | | |

| **Supplemental Table 3.** Effect of CR and exercise interventions on cognitive outcomes from linear mixed effects models additionally adjusted for BMI, peak VO_2_, gray matter volume, or cerebral blood flow at baseline and follow-up. | | | | |
| --- | --- | --- | --- | --- |
|  | **Change from baseline (95% CI)** | | |  |
|  | **Combined** | **CR+AT** | **CR+AT+RT** | **Regression coefficient of added covariate (95% CI)** |
| **Adjusted for BMI changes** |  |  |  |  |
| **MoCA^a^** | 1.65 (0.74, 2.56) | 1.83 (0.68, 2.98) | 1.47 (0.41, 2.53) | 0.06 (-0.10, 0.22) |
| **MoCA, *z*-score** | 0.70 (0.36, 1.04) | 0.77 (0.33, 1.21) | 0.62 (0.22, 1.03) | 0.04 (-0.02, 0.09) |
| **RAVLT^a^** |  |  |  |  |
| Immediate total | 0.56 (-0.08, 1.20) | 0.96 (0.11, 1.80) | 0.17 (-0.61, 0.95) | -0.007 (-0.10, 0.09) |
| Delayed total | 0.78 (-0.32, 1.88) | 1.25 (-0.13, 2.62) | 0.31 (-0.95, 1.57) | -0.03 (-0.24, 0.18) |
| **Cognitive domain *z*-scores** |  |  |  |  |
| Memory | 0.04 (-0.21, 0.29) | 0.16 (-0.17, 0.50) | -0.09 (-0.40, 0.23) | 0.005 (-0.03, 0.04) |
| Attention | 0.10 (-0.14, 0.33) | 0.03 (-0.28, 0.33) | 0.17 (-0.11, 0.45) | 0.02 (-0.02, 0.05) |
| Executive function | 0.04 (-0.21, 0.30) | 0.16 (-0.19, 0.52) | -0.08 (-0.40, 0.25) | -0.01 (-0.04, 0.02) |
| **Supplemental Table 3, continued.** | | | | |
| Visuospatial | -0.03 (-0.47, 0.40) | -0.10 (-0.72, 0.52) | 0.04 (-0.54, 0.61) | 0.0007 (-0.04, 0.04) |
| Language |  |  |  |  |
| Category Fluency | 0.13 (-0.09, 0.35) | 0.09 (-0.20, 0.37) | 0.17 (-0.09, 0.43) | -0.006 (-0.04, 0.03) |
| Phonemic Fluency | 0.20 (-0.09, 0.49) | -0.02 (-0.40, 0.36) | 0.42 (0.07, 0.76) | 0.003 (-0.04, 0.05) |
| **Adjusted for peak VO_2_ changes** |  |  |  |  |
| **MoCA^a^** | 1.56 (0.63, 2.49) | 1.65 (0.54, 2.76) | 1.47 (0.33, 2.60) | -0.03 (-0.27, 0.21) |
| **MoCA, *z*-score** | 0.61 (0.25, 0.97) | 0.65 (0.22, 1.08) | 0.58 (0.14, 1.02) | -0.01 (-0.10, 0.08) |
| **RAVLT^a^** |  |  |  |  |
| Immediate total | 0.34 (-0.34, 1.02) | 0.74 (-0.11, 1.59) | -0.06 (-0.91, 0.78) | 0.09 (-0.06, 0.25) |
| Delayed total | 0.84 (-0.31, 1.99) | 1.32 (-0.04, 2.68) | 0.37 (-1.03, 1.76) | 0.02 (-0.27, 0.31) |
| **Cognitive domain *z*-scores** |  |  |  |  |
| Memory | 0.13 (-0.14, 0.40) | 0.23 (-0.11, 0.58) | 0.03 (-0.31, 0.37) | -0.04 (-0.10, 0.03) |
| Attention | -0.002 (-0.25, 0.25) | -0.08 (-0.39, 0.24) | 0.07 (-0.24, 0.38) | 0.01 (-0.05, 0.08) |
| **Supplemental Table 3, continued.** | | | | |
| Executive function | 0.15 (-0.12, 0.42) | 0.25 (-0.11, 0.61) | 0.04 (-0.31, 0.39) | -0.02 (-0.07, 0.03) |
| Visuospatial | -0.05 (-0.50, 0.40) | -0.13 (-0.76, 0.49) | 0.03 (-0.56, 0.63) | 0.01 (-0.06, 0.09) |
| Language |  |  |  |  |
| Category Fluency | 0.18 (-0.05, 0.41) | 0.16 (-0.11, 0.43) | 0.20 (-0.08, 0.47) | -0.02 (-0.08, 0.04) |
| Phonemic Fluency | 0.28 (-0.28, 0.45) | 0.09 (-0.28, 0.45) | 0.47 (0.10, 0.83) | -0.05 (-0.12, 0.03) |
| **Adjusted for GM volume changes** |  |  |  |  |
| **MoCA^a^** | 1.46 (0.81, 2.11) | 1.62 (0.68, 2.55) | 1.30 (0.39, 2.21) | 0.09 (-0.20, 0.38) |
| **MoCA, *z*-score** | 0.59 (0.33, 0.85) | 0.66 (0.28, 1.03) | 0.52 (0.16, 0.88) | 0.03 (-0.09, 0.14) |
| **RAVLT^a^** |  |  |  |  |
| Immediate total | 0.55 (0.03, 1.07) | 0.89 (0.14, 1.64) | 0.21 (-0.52, 0.94) | 0.09 (-0.10, 0.30) |
| Delayed total | 0.80 (0.05, 1.56) | 1.31 (0.22, 2.40) | 0.30 (-0.75, 1.35) | -0.04 (-0.39, 0.38) |
| **Cognitive domain *z*-scores** |  |  |  |  |
| Memory | -0.02 (-0.23, 0.19) | 0.11 (-0.19, 0.41) | -0.15 (-0.44, 0.14) | -0.01 (-0.09, 0.08) |
| **Supplemental Table 3, continued.** | | | | |
| Attention | 0.01 (-0.17, 0.19) | -0.01 (-0.27, 0.25) | 0.04 (-0.21, 0.29) | -0.06 (-0.13, 0.02) |
| Executive function | 0.10 (-0.13, 0.33) | 0.23 (-0.11, 0.56) | -0.03 (-0.35, 0.30) | 0.003 (-0.07, 0.07) |
| Visuospatial | -0.05 (-0.45, 0.36) | -0.17 (-0.75, 0.41) | 0.07 (-0.49, 0.64) | 0.02 (-0.08, 0.13) |
| Language |  |  |  |  |
| Category Fluency | 0.16 (0.00, 0.31) | 0.13 (-0.10, 0.35) | 0.18 (-0.03, 0.40) | -0.02 (-0.10, 0.05) |
| Phonemic Fluency | 0.20 (-0.01, 0.42) | -0.04 (-0.35, 0.27) | 0.44 (0.15, 0.74) | 0.05 (-0.05, 0.14) |
| **Adjusted for GM CBF changes** |  |  |  |  |
| **MoCA^a^** | 1.50 (0.84, 2.15) | 1.71 (0.77, 2.65) | 1.29 (0.38, 2.20) | 0.03 (-0.03, 0.09) |
| **MoCA, *z*-score** | 0.60 (0.34, 0.86) | 0.69 (0.31, 1.06) | 0.51 (0.15, 0.87) | 0.01 (-0.01, 0.04) |
| **RAVLT^a^** |  |  |  |  |
| Immediate total | 0.57 (0.04, 1.09) | 0.93 (0.17, 1.68) | 0.21 (-0.52, 0.94) | 0.001 (-0.04, 0.05) |
| Delayed total | 0.80 (0.04, 1.57) | 1.31 (0.21, 2.41) | 0.30 (-0.76, 1.36) | 0.004 (-0.06, 0.08) |
| **Cognitive domain *z*-scores** |  |  |  |  |
| **Supplemental Table 3, continued.** | | | | |
| Memory | -0.01 (-0.22, 0.19) | 0.13 (-0.16, 0.43) | -0.16 (-0.45, 0.13) | 0.01 (-0.006, 0.03) |
| Attention | 0.00 (-0.18, 0.18) | -0.04 (-0.30, 0.22) | 0.04 (-0.21, 0.29) | -0.006 (-0.02, 0.01) |
| Executive function | 0.09 (-0.14, 0.33) | 0.21 (-0.13, 0.54) | -0.02 (-0.35, 0.31) | -0.01 (-0.03, 0.01) |
| Visuospatial | -0.02 (-0.40, 0.36) | -0.09 (-0.64, 0.46) | 0.05 (-0.48, 0.58) | 0.04 (0.01, 0.07) |
| Language |  |  |  |  |
| Category Fluency | 0.16 (0.004, 0.31) | 0.14 (-0.08, 0.36) | 0.18 (-0.04, 0.39) | 0.01 (-0.004, 0.03) |
| Phonemic Fluency | 0.22 (0.01, 0.43) | 0.01 (-0.30, 0.31) | 0.44 (0.14, 0.73) | 0.02 (-0.002, 0.04) |
| BMI, body mass index; CBF, cerebral blood flow; GM, gray matter; MoCA, Montreal Cognitive Assessment; RAVLT, Rey Auditory Verbal Learning Test; VO_2_, oxygen consumption rate | | | | |
| ^a^Also adjusted for age, sex, race, and years of education. | | | | |

**SUPPLEMENTAL METHODS**

***Exercise training***

Exercise sessions were conducted at the Wake Forest Clinical Research Center by Masters-level Certified Clinical Exercise physiologists with medical supervision. The CR+AT and CR+AT+RT groups met at different times to eliminate the potential for cross-group contamination. The AT prescription for each participant was based on initial evaluations of heart rate reserve (HRR), peak VO_2_, and Borg rating of perceived exertion. Intensity started at 40-50% of HRR and increased incrementally for the first 6-8 weeks until the participant could maintain 60-70% HRR for at least 20 minutes. Intensity was re-evaluated at least every 3 weeks by assessment of heart rate response during submaximal exercise. Depending on tolerance, AT duration progressed to 30 minutes performed continuously or in intermittent bouts. When a participant was able to complete 30 minutes of continuous AT, intensity was increased incrementally to 70-80% HRR. Periodic heart rate, rhythm, and blood pressure measures were performed to ensure safety and compliance.

Participants in the CR+AT+RT group had a short rest period before initiating the RT session. This intervention was designed to elicit skeletal muscle adaptations resulting in increased strength, mass, and quality. The RT regimen was in accordance with American Heart Association and American College of Sports Medicine guidelines for intensity, number of repetitions, number of sets, and frequency.^1,2^ All CR+AT+RT participants attended an orientation session where the correct use of equipment was demonstrated. Heart rate, rhythm, and blood pressure were monitored before and after each RT session. Participants exercised in small groups to allow rest and rotation between machines. Participants were instructed to complete the concentric phase of each movement in 2-3 seconds, pause briefly at the mid-point, and complete the eccentric phase in 2-3 seconds with 1-min rest intervals between sets. Amount of weight lifted, number of repetitions, and sets completed during each session were recorded. The RT regimen included 4 lower and 2 upper body exercises: leg curl (hamstrings), leg extension (quadriceps), leg press (knee extensors and hip flexors), calf raise (gastrocnemius), chest press (pectorals and triceps), and compound row (latissimus dorsi and biceps).

The RT prescription was based on a relative intensity level and progressed at a rate specific to each participant’s strength gain. The initial resistance setting on each machine was based on the participant’s 1-repetition maximum (1-RM) tested on the machine. During the first 3 weeks, resistance on each machine was 20-30% of participants’ 1-RM and 1 set of 8-12 repetitions were completed for each exercise. Starting at week 4, resistance increased to 40-50% of 1-RM and a second set of each exercise performed to volitional fatigue was added. For the remainder of the study, if >12 repetitions were completed on the second set in 2 consecutive sessions, resistance was increased for the next session. The 1-RM testing was repeated every 4 weeks to assess gains in strength and ensure optimal resistance settings, with a goal to achieve 70-80% 1-RM for the last 8 weeks of the study. To maintain a similar duration of exercise between groups, ~20 min of light chair-based range-of-motion, stretching, and flexibility exercises was included at the end of the 40-min AT sessions for the CR+AT group.

Multiple behavioral management strategies were employed to create a positive exercise environment and help maintain compliance. Participants who missed a session were promptly contacted to schedule make-up sessions, and all participants received individual counseling to discuss strategies to promote attendance and limit obstacles to participation. In the case of a change in health status or intermittent illness, modifications were made to the exercise prescription, including an evaluation of suitability to return to the intervention.

***MRI data processing***

High-resolution T1-weighted images were obtained with a magnetization prepared rapid gradient echo (MP-RAGE) sequence (TI = 900 ms; TR = 2300 ms; TE = 2.98 ms; 1 mm isotropic). Frontal lobe gray matter volume comprised the sum of non-overlapping major and minor gyri including: superior, middle, and inferior operculum; inferior triangularis; superior medial gyrus; supplementary motor area; superior, superior medial, middle, and inferior orbital; gyrus rectus; precentral gyrus; and the olfactory cortex. Temporal lobe gray matter volume included the bilateral sum of hippocampus; parahippocampus; amygdala; fusiform gyrus; Heschl’s gyrus; superior and middle temporal pole; and superior, middle, and inferior temporal gyri. T2-FLAIR images were obtained with a 3D inversion recovery sequence (TI = 1800 ms; TR = 5000 ms; TE = 383 ms; 1 mm isotropic). Arterial spin labeling (ASL) scans were performed with a whole brain multiphase pseudocontinuous (pc) sequence^3^ (tagging duration = 1.7 s, TI = 3 s, TR = 4 s, TE = 11 ms, reps = 81, FOV = 168x210 mm, 3x3x4 mm resolution, 36 axial slices, 8 averages with 8 images with unique phase offsets acquired in each), yielding a calibrated perfusion value for each voxel. MP-RAGE structural T1 and pcASL image files were converted to NIFTI format, and normalized, segmented, and corrected for head motion using CAT12 tools in SPM12. PCASL images were further corrected for partial volume and inconsistent volume filtering. Partial volume correction was used to generate gray and white matter CBF maps, which were registered onto T1 images.^4^ Image volumes with large motion artifacts or inconsistency in the PC-ASL time series were rejected before being averaged.^5^ The lesion growth algorithm for WMH volume segmentation was implemented in the LST toolbox v2.0.15.^6^

**Supplemental References**

1.	Williams MA, Haskell WL, Ades PA, et al. Resistance exercise in individuals with and

without cardiovascular disease: 2007 update: a scientific statement from the American Heart Association Council on Clinical Cardiology and Council on Nutrition, Physical Activity, and Metabolism. Circulation. 2007;116(5):572-584. doi:10.1161/CIRCULATIONAHA.107.185214
